# Flattening the curve before it flattens us: hospital critical care capacity limits and mortality from novel coronavirus (SARS-CoV2) cases in US counties

**DOI:** 10.1101/2020.04.01.20049759

**Authors:** Charles C. Branas, Andrew Rundle, Sen Pei, Wan Yang, Brendan G. Carr, Sarah Sims, Alexis Zebrowski, Ronan Doorley, Neil Schluger, James W. Quinn, Jeffrey Shaman

## Abstract

**Background:** As of March 26, 2020, the United States had the highest number of confirmed cases of Novel Coronavirus (COVID-19) of any country in the world. Hospital critical care is perhaps the most important medical system choke point in terms of preventing deaths in a disaster scenario such as the current COVID-19 pandemic. We therefore brought together previously established disease modeling estimates of the growth of the COVID-19 epidemic in the US under various social distancing contact reduction assumptions, with local estimates of the potential critical care surge response across all US counties.

**Methods:** Estimates of spatio-temporal COVID-19 demand and medical system critical care supply were calculated for all continental US counties. These estimates were statistically summarized and mapped for US counties, regions and urban versus non-urban areas. Estimates of COVID-19 infections and patients needing critical care were calculated from March 24, 2020 to April 24, 2020 for three different estimated population levels – 0%, 25%, and 50% – of contact reduction (through actions such as social distancing). Multiple national public and private datasets were linked and harmonized in order to calculate county-level critical care bed counts that included currently available beds and those that could be made available under four surge response scenarios – very low, low, medium, and high – as well as excess deaths stemming from inaccessible critical care.

**Results:** Surge response scenarios ranged from a very low total supply 77,588 critical care beds to a high total of 278,850 critical care beds. Over the four week study period, excess deaths from inaccessible critical care ranged from 24,688 in the very low response scenario to 13,268 in the high response scenario. Northeastern and urban counties were projected to be most affected by excess deaths due to critical care shortages, and counties in New York, Colorado, and Virginia were projected to exceed their critical care bed limits despite high levels of COVID-19 contact reduction. Over the four week study period, an estimated 12,203-19,594 excess deaths stemming from inaccessible critical care could be averted through greater preventive actions such as travel restrictions, publicly imposed contact precautions, greater availability of rapid testing for COVID-19, social distancing, self-isolation when sick, and similar interventions. An estimated 4,029-11,420 excess deaths stemming from inaccessible critical care could be averted through aggressive critical care surge response and preparations, including high clearance of ICU and non-ICU critical care beds and extraordinary measures like using a single ventilator for multiple patients.

**Conclusions:** Unless the epidemic curve of COVID-19 cases is flattened over an extended period of time, the US COVID-19 epidemic will cause a shortage of critical care beds and drive up otherwise preventable deaths. The findings here support value of preventive actions to flatten the epidemic curve, as well as the value of exercising extraordinary surge capacity measures to increase access to hospital critical care for severely ill COVID-19 patients.

## INTRODUCTION

The World Health Organization declared the novel coronavirus SARS-CoV2 a public health emergency of international concern on January 30, 2020 and a pandemic on March 12, 2020.^1^ Nations around the world are increasingly experiencing case clusters or community transmission. As of March 26, 2020, the United States had the highest number of confirmed cases of COVID-19, the disease caused by SARS-CoV2, of any country in the world.^2^

Multiple areas in the US are seeing dramatic increases in cases of COVID-19 and concerns are mounting those local medical system response capacities will be quickly exceeded. Hospital critical care is perhaps the most important medical system choke point in terms of preventing deaths in a disaster scenario such as with the current COVID-19 pandemic.^3,4^ A spectrum of critical care, from intensive care units to other serviceable hospital critical care structures, can be drafted in the event of mass disasters, potentially doubling hospital capacity in a crisis care surge situation.^5,6,7,8^

However, whether the nation’s potential hospital surge capacity is exactly double, or perhaps more or less than that, in the context of rapidly growing cases of COVID-19 in the US, remains unclear. We therefore brought together previously established disease modeling estimates^9^ of the growth of the COVID-19 epidemic in the US under various social distancing contact reduction assumptions, with local estimates of the potential critical care surge response^10^ across all US counties. Our objectives in doing this was to highlight US counties that are at risk of exceeding their critical care surge capacity limits within one month, indicate the typical time it would take these counties to exceed their critical care surge capacity limits, and estimate the excess mortality that would potentially result from exceeding critical care surge capacity limits in these counties. These objectives speak to the capabilities of the US medical system under disaster conditions and the usefulness of social distancing and other prevention strategies for slowing the presentation rate of severe COVID-19 cases to a point where the US critical care system can adapt in minimizing preventable mortality. ^11,12^

## METHODS

### Study setting and units of analysis

A full cohort of 3,108 US counties were included as our primary units of analysis. Aside from states, counties (equivalently known as parishes, boroughs, and independent cities in some states) are the major legally defined political and administrative units of the United States. As primary governmental divisions, county boundaries and names rarely change.^13^ Our county list included the District of Columbia as a county equivalent and we also tracked and accounted for any county names or Federal Information Processing Standards (FIPS) county codes that had changed over time across our various datasets. Alaska and Hawaii were not included in this analysis as these states were disconnected in the commuting dataset that was used to project the transmission and spatio-temporal spread of COVID-19 cases (these two states will be added in later models and as part of ongoing calculations).

All US counties were further aggregated into US regions and urban/non-urban classifications. US regions were defined using Census Bureau standards as Northeast, Midwest, South, and West.^14^ Counties were defined as urban or non-urban using the 2013 US Department of Agriculture rural-urban continuum classification (RUCC) scheme. Urban counties had RUCC codes 1-3 in this scheme and listed as metropolitan; non-urban counties had RUCC codes 4-9 and listed as non-metropolitan. This ordinal RUCC variable distinguishes counties by considering both their population size and proximity to metropolitan areas. In doing this it provides useful added information over and above simple categorizing of counties on the basis of population size, land area, proximity to metropolitan areas, or population density, as singular variables.^15,16^

Various estimated parameters of spatio-temporal COVID-19 demand and medical system critical care supply were then calculated for all continental US counties. These estimates were then statistically summarized and mapped for US counties, regions and urban versus non-urban areas. Geographic Information Systems software, ArcGIS Pro 2.5 (ESRI Inc., 2020, Redlands, CA), was used to manage analytic polygons and create maps.

### Estimates of spatio-temporal COVID-19 demand

A mathematical model was developed that simulates the spatiotemporal dynamics of infections. The details of this model are reported elsewhere.^17^ The model divides infections into two classes with separate rates of transmission: documented infected individuals and undocumented infected individuals. The spatial spread of COVID-19 is captured using Census Bureau commuting data to estimate the daily number of people traveling between counties and an estimated multiplicative factor.

Transmission dynamics were simulated for all US study counties over the period from February 21, 2020 to March 24, 2020 using an iterated filter-ensemble adjustment Kalman filter framework.^18,19,20^ This combined model-inference system estimated the trajectories of susceptible, exposed, documented infected, and undocumented infected populations in each county while simultaneously inferring model parameters for the average latent period, the average duration of infection, the transmission reduction factor for undocumented infections, the transmission rate for documented infections, the fraction of documented infections, and the previously mentioned travel multiplicative factor. To account for delays in infection confirmation, a time-to-event observation model using a Gamma distribution with a range of reporting delays and different maximum seeding was employed. Log-likelihood was used to identify the best fitting model-inference posterior.^8,16^

As in prior work^21^, the transmission of SARS-CoV2 under increasing reductions in population physical contact via control measures and behavior change was projected forward in time from March 24,2020 to April 24, 2020, using the optimized model parameter estimates. Control measures included travel restrictions between areas, self-quarantine and contact precautions that were publicly advocated or imposed, and greater availability of rapid testing for infection. Behavior changes in medical care-seeking due to increased awareness of COVID-19 and increased personal protective behavior (e.g., use of facemasks, social distancing, self-isolation when sick) were also considered. Three different contact reduction scenarios were projected, 0% (no contact reduction via controls and behavior change), 25% contact reduction, and 50% contact reduction.

### Estimates of spatio-temporal medical system critical care supply

Data on the counts and availability of various hospital beds that could be used for critical care were derived from the linkage and harmonization of different datasets for all US counties in the study. Several datasets, including four primary sources of data were used: (1) the 2020 Centers for Medicare & Medicaid Services (CMS), Health Care Information System (HCRIS) Data File, Sub-System Hospital Cost Report (CMS-2552-96 and CMS-2552-10), Section S-3, Part 1, Column 2; (2) the 2018 American Hospital Association (AHA) Annual Survey; (3) the 2020 US DHHS Health Resources and Services Administration, Area Health Resources Files (AHRF); and (4) the 2017-2019 CMS Medicare Provider of Services file, Medicare Cost Report, Hospital Compare Files.

The various types of hospital beds that could be used for critical care included intensive care unit (ICU) beds, as well as redirected operating room (OR) beds, post-anesthesia care unit (PACU) beds, and step-down beds. Critical care beds from all civilian US general medical-surgical, pediatric medical-surgical, and long-term acute care (LTAC) hospitals were included. No Veterans Affairs or military medical hospital facilities were included. Counts of ICU bed per hospital were summed as any reported: (a) general medical-surgical ICU beds, (b) surgical ICU beds, (c) coronary ICU beds, (d) burn care ICU beds, (e) pediatric ICU beds, and (f) other ICU beds. Neonatal ICU beds were excluded. Counts of ICU beds were the highest number of ICU beds reported by each US hospital across the four primary sources of data listed above.

A baseline critical care bed availability, in the absence of surge clearances, was established as 30% of existing ICU beds in each county being unoccupied and available. From this baseline, additional critical care bed counts were created for each county in our dataset that included beds that could be made available under four critical care surge response scenarios – very low, low, medium, and high (Table 1). Broadly, these four scenarios assumed the baseline that 30% of a hospital’s critical beds are unoccupied and available, that some currently occupied critical care beds can be cleared, that other specialized non-ICU beds can be redeployed as critical care beds, and, in the high scenario, that two critical care patients can be serviced using a single ventilator. In this way, existing critical care bed availability rates and occupied critical care bed clearance rates for purposes of meeting high-volume patient surges in disasters were incorporated into our estimates. Step-down bed counts were used where reported by hospitals in the four primary data sources; if hospitals did not report step-down beds, a 1:4 step-down-to-ICU bed ratio was assumed and ICU bed counts were multiplied by 1.25. One bed per OR was assumed. For hospitals that did not report PACU beds, a 1.5:1 PACU beds-to-OR ratio was assumed and ORs were multiplied by 1.5. One ventilator was assumed per critical care bed. The ability to put multiple patients on a single ventilator in order to meet demands in a high-volume disaster was also incorporated into our estimates.^22,23,24,25,26,27,28,29,30,31^

**Table 1.**
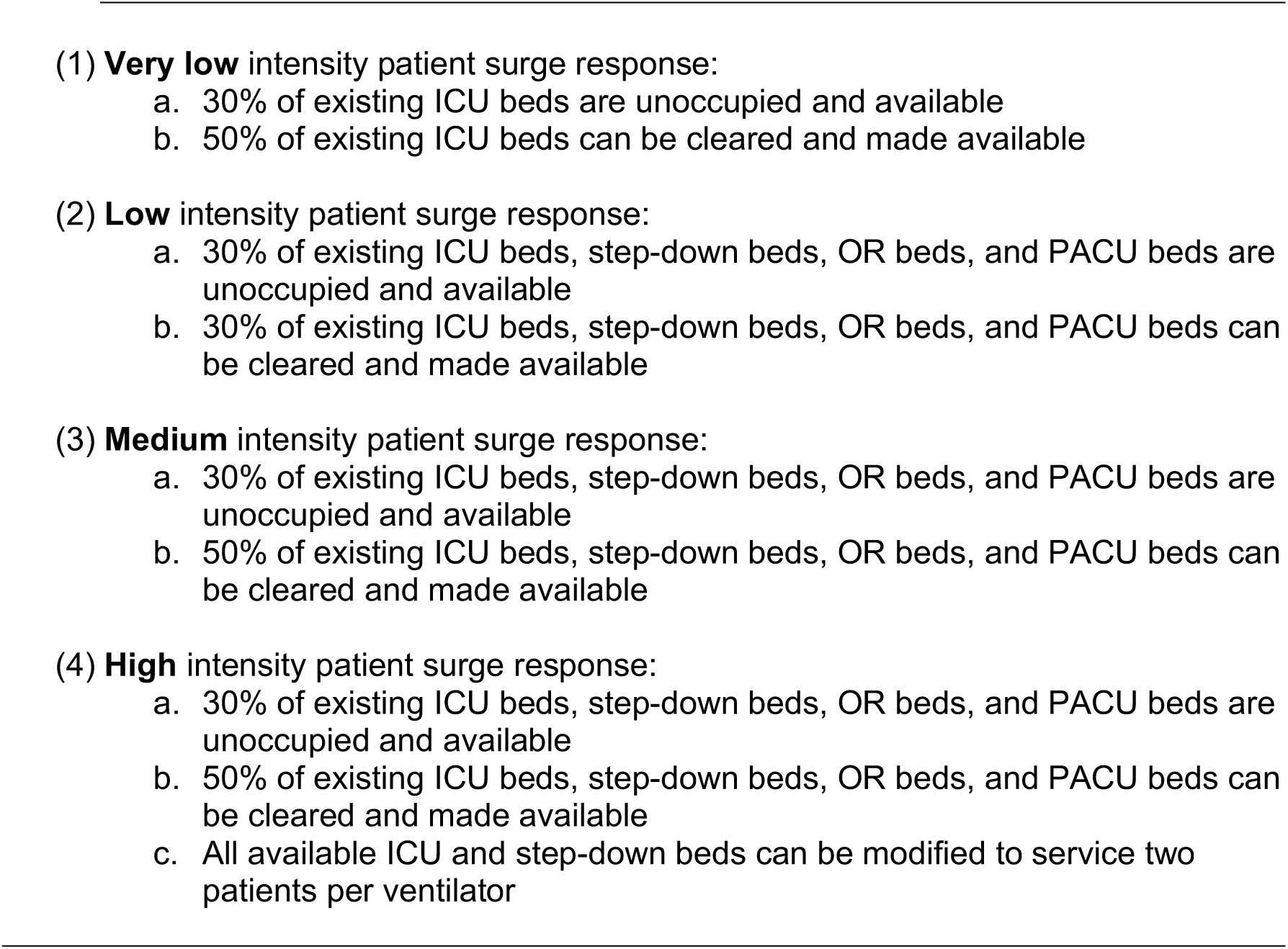
Calculated parameters underlying four critical care surge response scenarios.

Counties with zero beds were not included in calculating the mean numbers of days prior to exceeding critical care surge limits for each county. A typical ICU length of stay for COVID-19 patients was used to calculate the daily discharge rate from hospital critical care beds to recalculate critical care bed need for each day of the study period.^32^ Once a hospital’s critical care bed capacity was reached, patients who could not be admitted – i.e., new critical care bed need minus critical care bed discharges – were aggregated to calculate excess deaths due to lack of critical care access. Prior reports of the hospital course of care for COVID-19 patients showed that the vast majority of those admitted to the ICU were critical and only one-in-five of those who were critical survived, mostly because of ICU care. Thus, the percentage of critically ill patients that should have gone to the ICU but did not and survived should be much lower, likely only 5%; we therefore assumed a 95% mortality for patients that would have been placed in a critical care bed but did not because their local critical care bed capacity had been exceeded.^33^

## RESULTS

Of the US counties included in our analysis, 217 (7.0%) were in the Northeast, 1,055 (34.0%) were in the Midwest, 448 (14.4%) were in the South, and 1,388 (44.7%) were in the West. Additionally, 1,160 (37.3%) of the counties in our analysis were urban and 1,948 (62.7%) were non-urban.

The very low critical care surge response scenario had a total of 77,588 available critical beds, a mean +/-standard deviation (SD) of 24.0 +/-88.8 available critical beds per county, and a total of 24,688 excess deaths that would occur when critical care surge bed limits are exceeded over the projected 4 week study period. The low critical care surge response scenario had a total of 131,542 available critical beds, a mean +/-SD of 40.7 +/-143.1 available critical beds per county, and a total of 20,572 excess deaths that would occur when critical care surge bed limits are exceeded. The medium critical care surge response scenario had a total of 174,891 available critical beds, a mean +/-SD of 54.1 +/-190.9 available critical beds per county, and a total of 17,742 excess deaths that would occur when critical care surge bed limits are exceeded. The high critical care surge response scenario had a total of 278,850 available critical beds, a mean +/-SD of 86.2 +/-307.7 available critical beds per county, and a total of 13,268 excess deaths that would occur when critical care surge bed limits are exceeded.

When considering US regions, the number of counties with critical care beds exceeding their capacity within a month ranged from a high of 38 counties (17.5%) in the Northeast under the very low critical care surge response scenario with no contact reduction, to zero counties in multiple US regions under various scenario combinations. Urban counties were estimated to have greater numbers exceeding their critical care bed capacities within a month, from a maximum of 55 (4.7%) urban counties under the very low critical care surge response scenario with no contact reduction, to 6 (0.5%) urban counties under the high critical care surge response scenario with 50% contact reduction. (Table 2)

**Table 2.** Summary statistics of mean number of days prior to and total number of counties with critical care beds exceeding their bed limits within one month under different surge response and contact reduction scenarios, by US region and urbanicity

|  |  | Critical care surge response |  |  |  |
| --- | --- | --- | --- | --- | --- |
|  |  | Very low | Low | Medium | High |
| **Contact**<br>**reduction** | **0%** |  |  |  |  |
|  | Northeast | 21.0 days<br>(n=38 counties) | 22.4 days<br>(n=31 counties) | 22.5 days<br>(n=24 counties) | 23.5 days<br>(n=18 counties) |
|  | Midwest | 24.4 days<br>(n=5 counties) | 26.5 days<br>(n=4 counties) | 27.0 days<br>(n=3 counties) | 28.0 days<br>(n=1 counties) |
|  | South | 22.8 days<br>(n=11 counties) | 24.5 days<br>(n=11 counties) | 23.4 days<br>(n=8 counties) | 20.8 days<br>(n=5 counties) |
|  | West | 19.1 days<br>(n=10 counties) | 20.0 days<br>(n=6 counties) | 20.0 days<br>(n=5 counties) | 18.5 days<br>(n=2 counties) |
|  | Urban | 21.5 days<br>(n=55 counties) | 23.4 days<br>(n=48 counties) | 23.3 days<br>(n=36 counties) | 22.9 days<br>(n=23 counties) |
|  | Non-urban | 19.7 days<br>(n=9 counties) | 18.6 days<br>(n=5 counties) | 18.0 days<br>(n=4 counties) | 21.7 days<br>(n=3 counties) |
|  | **25%** |  |  |  |  |
|  | Northeast | 20.0 days<br>(n=27 counties) | 21.7 days<br>(n=20 counties) | 22.7 days<br>(n=17 counties) | 22.9 days<br>(n=9 counties) |
|  | Midwest | 25.0 days<br>(n=3 counties) | 28.0 days<br>(n=1 counties) | NA<br>(n=0 counties) | NA<br>(n=0 counties) |
|  | South | 22.7 days<br>(n=7 counties) | 21.5 days<br>(n=4 counties) | 21.5 days<br>(n=4 counties) | 21.5 days<br>(n=4 counties) |
|  | West | 15.3 days<br>(n=6 counties) | 17.8 days<br>(n=4 counties) | 16.3 days<br>(n=3 counties) | 13.5 days<br>(n=2 counties) |
|  | Urban | 20.6 days<br>(n=38 counties) | 21.9 days<br>(n=25 counties) | 22.5 days<br>(n=21 counties) | 22.5 days<br>(n=13 counties) |
|  | Non-urban | 16.2 days<br>(n=5 counties) | 17.8 days<br>(n=4 counties) | 16.3 days<br>(n=3 counties) | 13.5 days<br>(n=2 counties) |
|  | **50%** |  |  |  |  |
|  | Northeast | 18.8 days<br>(n=19 counties) | 21.6 days<br>(n=13 counties) | 20.4 days<br>(n=7 counties) | 22.2 days<br>(n=5 counties) |
|  | Midwest | 26 days<br>(n=1 counties) | NA<br>(n=0 counties) | NA<br>(n=0 counties) | NA<br>(n=0 counties) |
|  | South | 15.0 days<br>(n=2 counties) | 7.0 days<br>(n=1 counties) | 7.0 days<br>(n=1 counties) | 7.0 days<br>(n=1 counties) |
|  | West | 13.4 days<br>(n=5 counties) | 9.5 days<br>(n=2 counties) | 11.0 days<br>(n=2 counties) | 12.0 days<br>(n=1 counties) |
|  | Urban | 18.5 days<br>(n=23 counties) | 20.6 days<br>(n=14 counties) | 18.8 days<br>(n=8 counties) | 19.7 days<br>(n=6 counties) |
|  | Non-urban | 14.0 days<br>(n=4 counties) | 9.5 days<br>(n=2 counties) | 11.0 days<br>(n=2 counties) | 12.0 days<br>(n=1 counties) |

The 64 counties in the very low critical care surge response scenario with no contact reduction that were at risk of exceeding their bed limits were clustered in various locations across the US– a New York-New Jersey-Connecticut-northeastern Pennsylvania cluster, an eastern Massachusetts cluster, a southeastern Michigan cluster, a southeastern Louisiana cluster, a Colorado cluster, a Washington cluster, a Virginia cluster, and other dispersed counties in five other states. At the other extreme, the 7 counties in the high critical care surge response scenario with 50% contact reduction that were at risk of exceeding their bed limits were clustered in New York, Colorado, and Virginia. (Figure 1)

**Figure 1.**
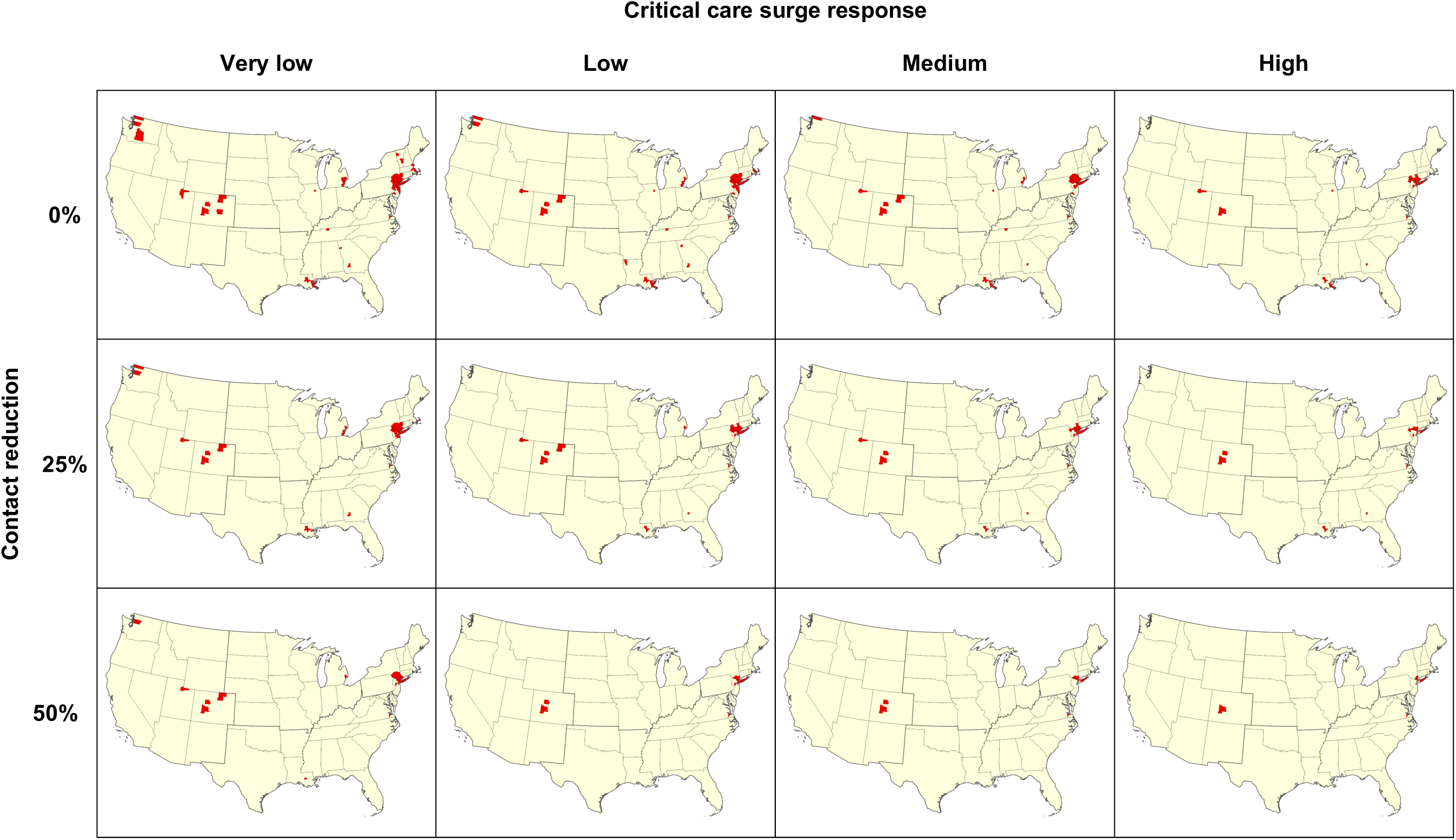
US counties exceeding critical care surge limits within one month (in red) under various surge response and contact reduction scenarios

The highest proportion of excess COVID-19 deaths that could have been averted with access to critical care was shown to occur in the Northeast US and urban counties over the month studied. As a measure of actions to flatten the epidemic curve, the difference in excess deaths between a 0% and a 50% contact reduction ranged from an estimated 12,203 to 19,594 excess deaths averted over a month. As a measure of the impact of aggressive critical care surge actions, the difference in excess deaths between the high and the very low critical care surge response scenarios ranged from an estimated 4,029 to 11,420 excess deaths averted over a month. As a measure of the impact of redeploying non-ICU beds for critical care surge response, the difference in excess deaths between the medium and the very low critical care surge response scenarios ranged from an estimated 3,050 to 6,946 excess deaths averted over a month. As a measure of the impact of putting two patients on a single ventilator, the difference in excess deaths between the high and the medium critical care surge response scenarios ranged from an estimated 979 to 4,474 excess deaths averted over a month. (Table 3)

**Table 3.** Summary statistics of *excess deaths* in counties that exceed critical care bed surge limits within one month under different surge response and contact reduction scenarios, by US region and urbanicity

|  |  | Critical care surge response |  |  |  |
| --- | --- | --- | --- | --- | --- |
|  |  | Very low | Low | Medium | High |
| **Contact**<br>**reduction** | **0%** |  |  |  |  |
|  | US Total | 24,688 | 20,572 | 17,742 | 13,268 |
|  | Northeast | 22,507 | 19,464 | 17,188 | 13,199 |
|  | Midwest | 1,537 | 839 | 435 | 1 |
|  | South | 289 | 152 | 41 | 29 |
|  | West | 355 | 117 | 78 | 39 |
|  | Urban | 24,554 | 20,494 | 17,685 | 13,228 |
|  | Non-urban | 134 | 78 | 57 | 40 |
|  | **25%** |  |  |  |  |
|  | US Total | 11,972 | 8,882 | 7,154 | 4,284 |
|  | Northeast | 11,329 | 8,829 | 7,124 | 4,261 |
|  | Midwest | 441 | 10 | 0 | 0 |
|  | South | 69 | 8 | 8 | 8 |
|  | West | 133 | 35 | 22 | 15 |
|  | Urban | 11,910 | 8,847 | 7,132 | 4,269 |
|  | Non-urban | 62 | 35 | 22 | 15 |
|  | **50%** |  |  |  |  |
|  | US Total | 5,094 | 3,082 | 2,044 | 1,065 |
|  | Northeast | 5,022 | 3,069 | 2,033 | 1,055 |
|  | Midwest | 11 | 0 | 0 | 0 |
|  | South | 7 | 2 | 2 | 2 |
|  | West | 54 | 11 | 9 | 8 |
|  | Urban | 5,063 | 3,071 | 2,035 | 1,057 |
|  | Non-urban | 31 | 11 | 9 | 8 |

The increase in critical care beds that could be achieved under the various surge response scenarios was highly correlated with the number of beds estimated under the baseline critical care bed availability model. Focusing on the medium critical care surge capacity scenario, the gain in critical care beds under this scenario was highly correlated with the estimated beds available under baseline critical care bed availability (r=0.97). Regression analyses found that for each baseline critical care bed, 4.61 (95% CI 4.57, 4.65) additional critical care beds could be gained under the medium critical care surge capacity scenario. The counties that could generate the largest gains in beds under these surge capacity scenarios were counties that already had substantial hospital infrastructure and these counties were typically large urban areas.

## DISCUSSION

An inversely proportional relationship is evident between available critical care beds and excess deaths that would occur when critical care surge bed limits are exceeded. The value of “flattening the curve” – that is, the difference between having none and achieving a 50% contact reduction – is potentially sizeable in terms of affording the US medical system, especially the choke point of hospital critical care, the necessary time to prepare and be able to handle a manageable throughput volume of severely ill COVID-19 cases.

Roughly 10,000 to 20,000 excess deaths stemming from inaccessible critical care could be averted within the 4 week study period through greater preventive actions such as travel restrictions, publicly imposed contact precautions, greater availability of rapid testing for COVID-19, social distancing, self-isolation when sick, and similar interventions. Moreover, roughly 4,000 to 11,000 excess deaths stemming from inaccessible critical care could be averted within the 4 week study period through aggressive critical care surge response and preparations, including high clearance of ICU and non-ICU critical care beds and extraordinary measures like using a single ventilator for multiple patients. Adding in the capability of putting two patients on a single ventilator in order to meet demands in a high-volume disaster such as the current pandemic could save the lives of an additional 979 to 4,474 critically ill COVID-19 patients over a month.^34^

The highest proportion of excess COVID-19 deaths that could have been averted with access to critical care are shown to occur in the Northeast US and urban counties over the projected month studied, a reflection of the COVID-19 caseload in the New York City region. However, seven other major clusters of potentially preventable deaths due inadequate critical care access were estimated across the US. Assuming a high level of contact reduction and a high level of critical care surge response, these clusters most prominently include counties in New York, Colorado, and Virginia, although relaxations of these high assumptions were shown to add clusters in states such as Louisiana, Michigan, and Virginia.

While large urban areas generally had the largest capacity to generate additional critical care beds under our surge capacity models, these same urban counties currently, or in the near future, are predicted to have the largest numbers of COVID-19 cases. A major concern that then arises is whether the critical care surge capacity in these counties is sufficient to care for the projected numbers of COVID-19 cases that will continue to mount. As a matching concern, the relocation or travel of urban residents with undetected COVID-19 infection to non-urban areas that appear to be relatively unaffected may overwhelm the relatively limited critical care capacity in otherwise isolated non-urban regions. A potential example of this in the current dataset is the Colorado cluster that includes major winter vacation resorts that may have had visitors in from major cities in the US and internationally soon before public notification of the current COVID-19 crisis.

The estimates presented here are based on long-established federal and professional agency inventories and estimations of hospitals and hospital beds across the US. However, a major limitation is that the models presented here use data on physical infrastructure but do not account for staffing or ventilator supplies. Healthcare workers, especially those involved in critical care, are at high risk for COVID-19 infection and thus there may be staffing shortages that reduce the utility of the critical care beds that could be gained under surge responses. There have already been reports of hospitals being unable to accept patients, not because of lack of beds but due to lack of staff to cover those beds. Some states, like New York, are currently recruiting retired healthcare workers to assist with staffing shortfalls, an approach that might be generally applicable in alleviating shortfalls during the current epidemic. These retirees are, however, generally older and can be particularly vulnerable to poor outcomes from COVID-19. Our models also cannot account for the innovation, ingenuity and perseverance of medical staff, many of whom are trained to work in crisis situations. It is likely that medical staff will find solutions that are unanticipated by our models, that can subsequently be included as they become known and more widely applied across healthcare systems. Future analyses should also incorporate counts of ventilators in addition to critical care beds. Our models also did not account for heterogeneities arising from specific high-risk communities in different localities. For instance, places with large elderly populations or high levels of pre-existing respiratory, cardiovascular, or immunocompromised conditions would have even higher mortality rates. Future analyses could also account for underlying population risk factors such as these.

As has been often discussed, unless the epidemic curve of COVID-19 cases is flattened over an extended period of time, the global pandemic, and the US COVID-19 epidemic which is now the world’s largest, will cause a shortage of critical care beds and drive up otherwise preventable deaths.^35^ Despite this ominous forecast, the current paper has demonstrated the potential value of preventive actions to flatten the epidemic curve, as well as the value of exercising extraordinary surge capacity measures to increase access to hospital critical care for severely ill COVID-19 patients.

## Data Availability

Data available upon request and with appropriate and complete attribution of scientific author group.

## References

1 Kluge HHP. WHO announces COVID-19 outbreak a pandemic. March 12, 2020. URL: http://www.euro.who.int/en/health-topics/health-emergencies/coronavirus-covid-19/news/news/2020/3/who-announces-covid-19-outbreak-a-pandemic. Last accessed: March 29, 2020.

2 Soucheray S. US COVID-19 cases surge past 82,000, highest total in world. University of Minnesota Center for Infectious Disease Research and Policy. March 26, 2020. URL: http://www.cidrap.umn.edu/news-perspective/2020/03/us-covid-19-cases-surge-past-82000-highest-total-world. Last accessed: March 29, 2020.

3 Carr, B. G., Addyson, D. K., & Kahn, J. M. (2010). Variation in critical care beds per capita in the United States: implications for pandemic and disaster planning. JAMA, Journal of the American Medical Association, 303(14), 1371–1372.

4 Carr, B.G., Walsh, L., Williams, J.C., Pryor, J.P. and Branas, C.C., 2016. A geographic simulation model for the treatment of trauma patients in disasters. Prehospital and Disaster Medicine, 31(4), 413–421.

5 Hossain, T., Ghazipura, M., & Dichter, J. R. (2019). Intensive Care Role in Disaster Management Critical Care Clinics. Critical Care Clinics, 35(4), 535–550.

6 Corcoran, S. P., Niven, A. S., & Reese, J. M. (2012). Critical care management of major disasters: a practical guide to disaster preparation in the intensive care unit. Journal of Intensive Care Medicine, 27(1), 3–10.

7 Society for Critical Care Medicine. U.S. ICU Resource Availability for COVID-19. Version 2, 3/19/2020 1:49pm

8 Hick, J.L., Einav, S., Hanfling, D., Kissoon, N., Dichter, J.R., Devereaux, A.V., Christian, M.D. and Task Force for Mass Critical Care, 2014. Surge capacity principles: care of the critically ill and injured during pandemics and disasters: CHEST consensus statement. Chest, 146(4), e1S–e16S.

9 Li R, Pei S, Chen B, Song Y, Zhang T, Yang W, Shaman J. Substantial undocumented infection facilitates the rapid dissemination of novel coronavirus (COVID-19). Science. 2020 Mar 16.

10 Carr, B.G., Walsh, L., Williams, J.C., Pryor, J.P. and Branas, C.C., 2016. A geographic simulation model for the treatment of trauma patients in disasters. Prehospital and disaster medicine, 31(4), 413–421.

11 Emanuel, Ezekiel J., Govind Persad, Ross Upshur, Beatriz Thome, Michael Parker, Aaron Glickman, Cathy Zhang, Connor Boyle, Maxwell Smith, and James P. Phillips. “Fair allocation of scarce medical resources in the time of Covid-19.” NEJM (2020).

12 Klarevas L, Rajan S, Branas C, Keyes K. Experts chide Trump on coronavirus, the economy and depression. New York Daily News. March 25, 2020.

13 Geographic Areas Reference Manual. Washington (DC): Census Bureau (US); 2001. Available at: http://www.census.gov/geo/www/garm.html. Accessed September 2, 2004.

14 US Census Bureau. Census Regions and Divisions of the Unites States. URL: https://www2.census.gov/geo/pdfs/maps-data/maps/reference/us_regdiv.pdf. Last accessed: March 29, 2020.

15 Cromartie J. United States Department of Agriculture, Economic Research Service. Rural-Urban Continuum Codes Documentation. 2019. URL: https://www.ers.usda.gov/data-products/rural-urban-continuum-codes/documentation/. Last accessed: March 30, 2020.

16 Branas CC, Nance ML, Elliott MR, Richmond TS, Schwab CW. Urban–rural shifts in intentional firearm death: different causes, same results. American journal of public health. 2004 Oct;94(10):1750–5.

17 Pei S, Shaman J. Simulation of SARS-CoV2 Spread and Intervention Effects in the Continental US with Variable Contact Rates, March 24 2020. March 26, 2020. Available at https://github.com/shaman-lab/COVID-19Projection_0324.

18 E. L. Ionides, C. Bretó, A. A. King, Inference for nonlinear dynamical systems. Proc. Natl. Acad. Sci. U.S.A. 103, 18438–18443 (2006).

19 A. A. King, E. L. Ionides, M. Pascual, M. J. Bouma, Inapparent infections and cholera dynamics. Nature 454, 877–880 (2008).

20 S. Pei, F. Morone, F. Liljeros, H. Makse, J. L. Shaman, Inference and control of the nosocomial transmission of methicillin-resistant Staphylococcus aureus. eLife 7, e40977 (2018).

21 Glanz J, Leatherby L, Bloch M, Smith M, Buchanan L, Wu J, Bogel-Burroughs N. Coronavirus Could Overwhelm U.S. Without Urgent Action, Estimates Say. New York Times, March 20, 2020. URL: https://www.nytimes.com/interactive/2020/03/20/us/coronavirus-model-us-outbreak.html. Last accessed: March 31, 2020.

22 Hossain, T., Ghazipura, M., & Dichter, J. R. (2019). Intensive Care Role in Disaster Management Critical Care Clinics. Critical Care Clinics, 35(4), 535–550.

23 Corcoran, S. P., Niven, A. S., & Reese, J. M. (2012). Critical care management of major disasters: a practical guide to disaster preparation in the intensive care unit. Journal of intensive care medicine, 27(1), 3–10.

24 Marcon, E., Kharraja, S., Smolski, N., Luquet, B. and Viale, J.P., 2003. Determining the number of beds in the postanesthesia care unit: a computer simulation flow approach. Anesthesia & Analgesia, 96(5), 1415–1423.

25 Neyman, G., & Irvin, C. B. (2006). A single ventilator for multiple simulated patients to meet disaster surge. Academic Emergency Medicine, 13(11), 1246–1249.

26 Hossain, T., Ghazipura, M., & Dichter, J. R. (2019). Intensive Care Role in Disaster Management Critical Care Clinics. Critical Care Clinics, 35(4), 535–550.

27 Corcoran, S. P., Niven, A. S., & Reese, J. M. (2012). Critical care management of major disasters: a practical guide to disaster preparation in the intensive care unit. Journal of Intensive Care Medicine, 27(1), 3–10.

28 Society for Critical Care Medicine. U.S. ICU Resource Availability for COVID-19. Version 2, 3/19/2020 1:49pm

29 Hick, J.L., Einav, S., Hanfling, D., Kissoon, N., Dichter, J.R., Devereaux, A.V., Christian, M.D. and Task Force for Mass Critical Care, 2014. Surge capacity principles: care of the critically ill and injured during pandemics and disasters: CHEST consensus statement. Chest, 146(4), e1S–e16S.

30 Carr, B.G., Walsh, L., Williams, J.C., Pryor, J.P. and Branas, C.C., 2016. A geographic simulation model for the treatment of trauma patients in disasters. Prehospital and Disaster Medicine, 31(4), 413–421.

31 Giroir BP, Adama J. Optimizing ventilator use during the COVID-19 pandemic. US Public Health Service Commissioned Corps. March 31, 2020.

32 Pei S, Shaman J. Simulation of SARS-CoV2 Spread and Intervention Effects in the Continental US with Variable Contact Rates, March 24 2020. March 26, 2020. Available at https://github.com/shaman-lab/COVID-19Projection_0324.

33 Zhou, Fei, Ting Yu, Ronghui Du, Guohui Fan, Ying Liu, Zhibo Liu, Jie Xiang et al. Clinical course and risk factors for mortality of adult inpatients with COVID-19 in Wuhan, China: a retrospective cohort study. Lancet (2020).

34 Giroir BP, Adama J. Optimizing ventilator use during the COVID-19 pandemic. US Public Health Service Commissioned Corps. March 31, 2020.

35 Emanuel, Ezekiel J., Govind Persad, Ross Upshur, Beatriz Thome, Michael Parker, Aaron Glickman, Cathy Zhang, Connor Boyle, Maxwell Smith, and James P. Phillips. Fair allocation of scarce medical resources in the time of Covid-19. New England Journal of Medicine (2020).

